# Migration, Infectious Diseases and Drug Addiction in Russia

**DOI:** 10.1101/2020.10.09.20209791

**Authors:** Marina Lifshits, Natalia Neklyudova

## Abstract

The aim of the paper is to analyze the possible impact of various aspects of internal and external migration in Russia’s regions on the prevalence of the following social dangerous diseases: HIV, active tuberculosis, syphilis, drug addiction, acute and chronic viral hepatitis *B* and *C*. We analyzed the papers that concern the impact of migration on the health of the host territory population. The main research methods are econometric and correlation analysis. We constructed panel models for each disease. The models tested various socioeconomic indicators (including education level, cash income, housing improvements and the incidence of alcoholism), as well as climatic, geographical and demographic indicators. Influence of disease incidence in the neighbouring regions is also considered. Five various indicators of migration were tested. They characterize labour immigration, internal and external migration inflows to the regions and share of people born outside the region. We tried to track changes of factors that influence spread of diseases over time. It allowed us to correct the conclusions drawn earlier. In the course of the research, positive significant statistical correlation of the following indicators of migration and disease incidence was established:

1. foreign citizens employment and incidence of syphilis in 2005;
2. share of internal migrants and incidence of drug addiction in 2005;
3. foreign citizens employment and incidence of drug addiction in 2006-2016;
4. foreign citizens inflow and detection of chronic viral hepatitis in 2010;
5. foreign citizens inflow and detection of acute hepatitis *C* in 2011-2016.

**JEL F22; I1; R23**

## 1. Introduction

Historically spatial movements of people played an important role in the spread of socially dangerous diseases, especially infectious ones. In the modern world, which learned to apply precautionary measures against many dangers, the situation is not so unambiguous and needs research. Migration raises many socioeconomic issues, including the impact of migration on the health of the host society and migrants themselves. Health issues faced by migrants are often similar to those faced by the rest of the population, although in some cases they may be specific. However, the link between migration and the importation of contagious diseases, as well as the impact of the presence of migrants on their spread (including “social dangerous” diseases), have not been fully studied yet.

Some researchers have found that migrants who moved less than two years ago were healthier at the time of their arrival (except when they were diagnosed with contagious diseases) than local residents of the host country. This phenomenon, called “healthy migrant effect”, can be explained by the self-selection of healthier people who decide to move from their homeland to another country. This effect was confirmed by studies in the US (Singh et al., 2001), Canada (Hyman, 2001), Australia (Armfield et al., 2000), and several countries in Western Europe (Toma, 2001). However, “healthy migrant effect” tends to die out. Some time after migrating, the health of migrants and aborigines in the host country is usually compared (Newbold, 2005). However, sometimes it does not happen. For example, the overall mortality rates of immigrants from Latin America are lower than that of the indigenous people of the United States, despite their less favorable socioeconomic conditions (Markides et al., 2005). A similar result was obtained in studies of Morocco citizens in France (Khlat et al., 2003) and the Turkish population in Germany (Razum, 1998).

Another explanation of better health of migrants in comparison with aboriginals of host countries was called “salmon effect”. As many salmons come back from the ocean to natal rivers, where they once began their lives to spawn and die, alike the elderly quite often want to return home. Since old and gravely ill migrants often return to their homeland, the average expected life expectancy of those who remain is increased. In any case, this is exactly what happens in the US (Abraido-Lanza et al., 1999; Franzini et al., 2001; Turra et al., 2008). The research conducted in China confirms existence of this phenomenon concerning internal migration (Lu et al., 2014).

Kislitsyna (2013) after analysing data from the European Social Survey and the Russian Monitoring of Economic Situation and Health, identified statistically significant differences in self-reported health of migrants and indigenous people in 14 out of 29 countries, including Russia. She notes that most countries lack statistics to compare the health of migrants and indigenous people. This is particularly the case of Russia.

As for the possible role of migration in the importation of contagious diseases in the host community, the WHO regional office for Europe insists, “there is no systematic link between these phenomena. Communicable diseases are associated primarily with poverty. The European Region has a long experience of communicable diseases such as tuberculosis, HIV/AIDS, hepatitis, measles and rubella and has significantly reduced their burden during economic development, through better housing conditions, access to safe water, adequate sanitation, efficient health systems and access to vaccines and antibiotics. These diseases have not, however, been eliminated and still exist in the European Region, independently of migration. … The risk for importation of exotic and rare infectious agents into Europe …, is extremely low. Experience has shown that, when importation occurs, it involves regular travelers, tourists or health care workers rather than refugees or migrants”.^1^

At the same time, the Infectious Disease Epidemiology Annual Report published in Robert Koch Institute (Beermann S. et al., 2017), points to the negative consequences of a mass influx of migrants for the health of the German population. The report lists diseases whose prevalence has increased including: cholera, hepatitis, AIDS, leprosy, malaria, syphilis, and tuberculosis.

Most European studies confirm that the level of HIV-associated tuberculosis among migrants is higher than among the local population (Tavares et al., 2017).

A number of European researchers mark the underreporting of cases. A lesser degree of underreporting of infectious diseases in reports among migrants is noted compared to the underreporting of illnesses among the local population. Some authors see the reason that the share of unreported cases of tuberculosis is less among migrants than that of the local population is in the strengthened communicable disease surveillance over migrants (Melosini, 2012; Farchi et al., 2008). Nightingale et al. assumed that a lower degree of underreporting of hepatitis *B* and *C* among migrants compared to local residents is associated with mandatory screening of arriving migrants (Nightingale et al., 2009).

In Russia the situation is similar. All foreign citizens are subject to compulsory medical examination on syphilis, tuberculosis and HIV infection, unlike the citizens of the Russian Federation, and this fact can be reflected in incidence indicators. For example, the incidence of HIV, tuberculosis and syphilis among foreign citizens was 1.5 times higher than that of local residents in 2010–2013 (Shcherbak and Ulyukin, 2014). According to data of the Central Research Institute of Tuberculosis, among every 100,000 examined Russians and every 100,000 thousand foreign citizens in Russia, tuberculosis is detected 2.65 times less often among the former, but HIV - 2.93 times more often than among the latter. Such distinctions are connected with the fact that incidence of tuberculosis is higher in the majority of the countries of origin, than in Russia, while the prevalence of HIV infection is lower (Demikhova and Nechaeva, 2016).

Prokhorov (2009) believed that the main factor affecting people’s health is the socioeconomic situation status of individual population groups. It is obvious that this can affect the level of migrant morbidity. Rashkevich et al. (2016) note a rate of latent (3 times) and active tuberculosis (10 times) among children from migrant families in Smolensk Oblast in 2014– 2015. Migrant children are more often in contact with tuberculosis patients (6 times more often than children permanently residing in Smolensk Oblast) and 25% less likely to have BCG-vectored vaccination. The study by Maltseva et al. (2009) indicated a fairly high level of hepatitis infection among foreign citizens who arrived in Khabarovsk with working visa. At the same time, 17% of surveyed migrants possess antibodies to hepatitis *E* not endemic in Khabarovsk Krai. Alsalikh et al. (2017) came to a conclusion that a significant share of labour migrants arriving in Russia is infected with hepatitis *C*, which suggests a high probability of the importation of this infection. Istomin et al. (2017) conclude that migration significantly affects the spread of HIV. Solonin et al. (2010) speak of a significant level of hepatitis *E* infection among citizens coming from China to the territory of the Russian Federation. Struin and Shubina (2015) analysed studies (mainly foreign) on the incidence of social infections among migrants in various countries; the analysis also confirms that the incidence of syphilis, HIV and other sexually transmitted diseases among migrants is higher than among local residents.

Thus, the majority of researchers engaged in studying the impact of foreign citizens on the epidemiological situation in the host territory agree that migration can make a significant contribution to the spread of socially dangerous diseases, contrary to the statements of WHO/Europe. Nowadays, the impact of international migration on the spread of socially dangerous diseases remains debatable.

The present paper aims to use probability models to assess the possible impact of different aspects of migration on the spread of a number of socially dangerous diseases in the Russian regions. To achieve this, we set the objective to perform an econometric analysis of the spread of diseases that can be theoretically linked with migration. This is drug addiction and some diseases from the List of socially dangerous diseases,^2^ for which there is enough statistics for modelling.

We used data from Rosstat, Unified Interdepartmental Statistical Information System (EMISS)^3^ and Ministry of Health of the Russian Federation.

## 2. Data and methodology

We selected the following diseases for the purposes of our study: HIV (**D1**); active tuberculosis^4^ (**D2**); syphilis (**D3**); drug addiction (**D4**); chronic viral hepatitis^5^ (**D5**); acute viral hepatitis *B* (**D6**) and acute viral hepatitis *C* (**D7**). The EMISS database contains statistics of the Ministry of Health in the regional context since 2005 for D1-D4 and for other diseases just since 2010. In general, the situation in regions differs in a big variety.

The map of Federal districts of the Russian Federation is presented in Figure 1. The smallest incidence of HIV infection, syphilis, chronic and acute viral hepatitis was observed mainly in the North Caucasian Federal District (NCFD), of drug addiction – in the Southern Federal District (SFD), of tuberculosis – in the Central Federal District (CFD) and NCFD. The greatest incidence of HIV infection was determined in the Ural Federal District (UFD) and the Siberian Federal District (SibFD); of tuberculosis – in the Far Eastern Federal District (FEFD); of syphilis – in the FEFD and the SibFD. The largest incidence of drug addiction and acute viral hepatitis *C* was seen in the UFD; of chronic viral hepatitis – in the Northwestern Federal District (NWFD); of acute viral hepatitis *B* – in the CFD and the NWFD.

**Fig. 1:**
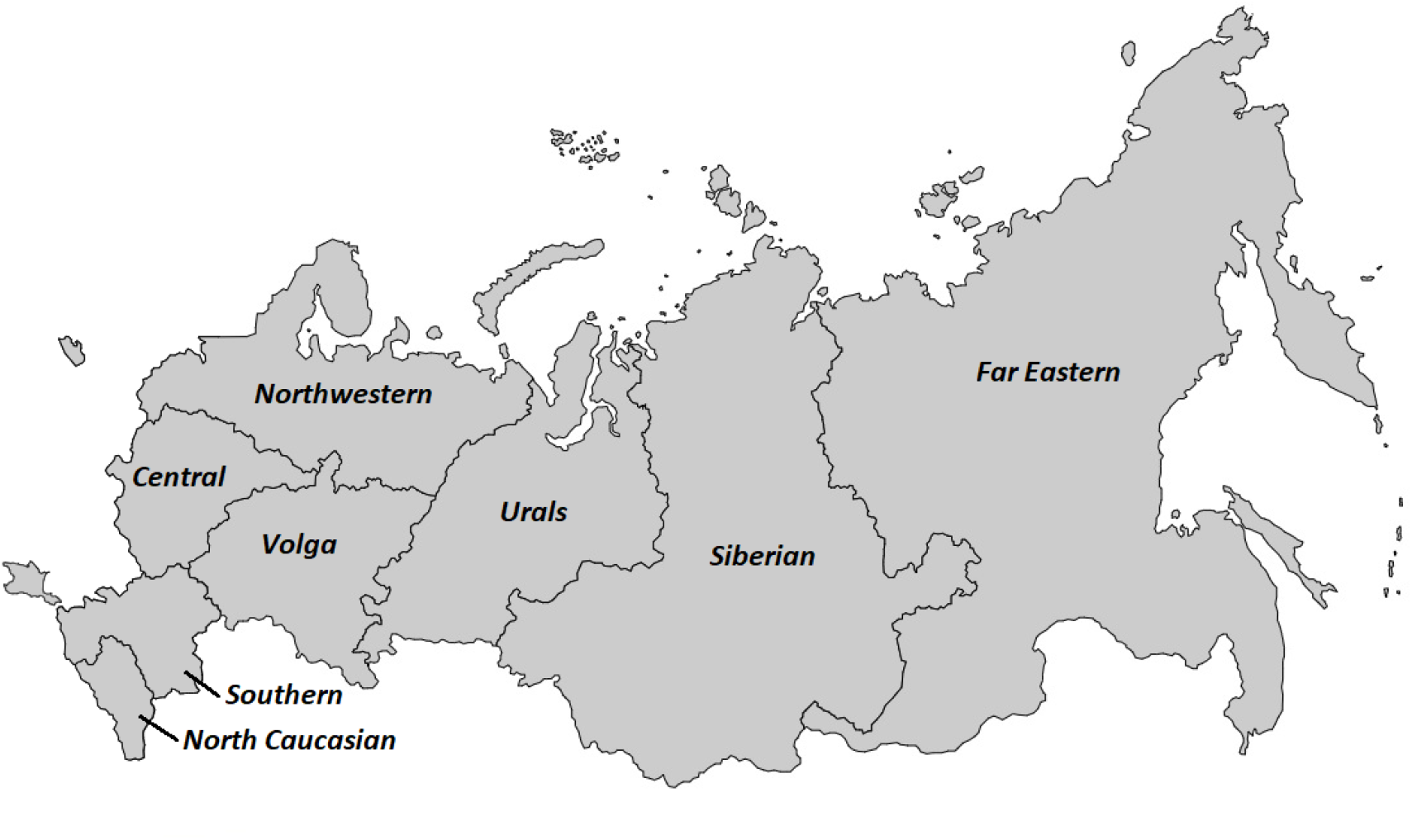
Federal districts of the Russian Federation

Table 1 lists Federal Districts with the highest and the lowest incidence of each of the socially dangerous diseases under review. At the first reference to the region, we indicate Federal district to which the region refers. There is no region of the Russian Federation that would be included in the top ten or the worst in each of the considered diseases.

**Table 1:** The highest and lowest incidence of socially dangerous diseases (per 100,000 people) in the Russian Federation regions

| *Regions and years with the highest incidence* | *Regions and years with the lowest incidence* |
| --- | --- |
| **D1. HIV (2005-2016)** |  |

<sup>2</sup> Approved by the Resolution of the Government of the Russian Federation no. 715, dated 01.12.2004. Available at: http://base.garant.ru/12137881/ (accessed: 19.03.2018).
<sup>3</sup> Unified Interdepartmental Statistical Information System. Available at: https://fedstat.ru/organizations/ (accessed: 01.07.2019).
<sup>4</sup> The Ministry of Health provides statistics separately on “active tuberculosis” and “tuberculosis of respiratory tract”. In the List of socially significant diseases at Rosstat website only “active tuberculosis” is listed, although the List approved by the Government of the Russian Federation contains “tuberculosis” without specification.
<sup>5</sup> diagnosed for the first time; the virus is not specified (hepatitis *B* and *C* can become chronic)

|  |  |
| --- | --- |
| Kemerovo Oblast (SibFD)<br>2014 – 240, 2015 – 238, 2012 – 219 | Nenets Autonomous Okrug (NWFD) 2015 – 0.0;<br>Rep. of Tyva (SibFD) 2006 – 0.0, 2008 – 1.3 |
| **D2. Active tuberculosis (2005-2016)** |  |
| Rep. of Tyva 2005 – 250, 2006 – 246, 2008 – 240 | Nenets Autonomous Okrug<br>2010-2011 – 0.0, 2016 – 20.5 |
| **D3. Syphilis (2005-2016)** |  |
| Chukotka Autonomous Okrug (FEFD) 2006 – 663;<br>Rep. of Tyva 2005 – 504, 2008 – 487 | Nenets Autonomous Okrug 2013 – 2.3;<br>Rep. of Dagestan (NCFD) 2016 – 3.5, 2015 – 3.7 |
| **D4. Drug addiction (2005-2016)** |  |
| Jewish Autonomous Oblast (FEFD) 2016 – 66.8;<br>Irkutsk Oblast (SibFD) 2005 – 56.7;<br>Kemerovo Oblast 2006 – 52.9 | Chukotka Autonomous Okrug 2010, 2012, 2014-2016 – 0.0<br>Nenets Autonomous Okrug 2016 – 0.0 |
| **D5. Chronic viral hepatitis (2010-2016)** |  |
| Kamchatka Krai (FEFD) 2010 – 233, 2011 – 218,<br>2012 – 204 | Chechen Rep. 2015 – 0.8, 2014 – 1.3, 2016 – 2.2 |
| **D6. Acute viral hepatitis B (2010-2017)** |  |
| Chukotka Autonomous Okrug 2010 – 7.84;<br>Tomsk Oblast (SibFD) 2013 – 5.90, 2014 – 5.58 | There is zero value in 20 regions of all Federal Districts |
| **D7. Acute viral hepatitis C (2010-2017)** |  |
| Rep. of Karelia (NWFD) 2010 – 8.24;<br>Magadan Oblast (FEFD) 2011 – 6.45;<br>Kurgan Oblast (UFD) 2011 – 5.47 | There is zero value in 12 regions of all Federal Districts, except for Central one. |

*Source: EMISS data*

Statistical data testify that the incidence of tuberculosis, syphilis and acute viral hepatitis *B* and *C* obviously declines while the HIV infection epidemic tends to grow.

In 2010 HIV infection took the 4th place on number of the diseased in the Russian Federation among the seven considered diseases (after tuberculosis, syphilis and chronic viral hepatitis), but since 2014 it has taken the first place. In 2016 the number of people diagnosed with HIV for the first time reached 84,000 cases. It was followed by active tuberculosis (77,000), chronic viral hepatitis (68,000), syphilis (31,000), drug abuse (16,000), hepatitis *C* (1,806) and hepatitis *B* (1,378).

On each of the considered diseases, there are annual cases of diseases almost in each region of the Russian Federation, and in general there are enough data to construct probability panel models for disease dissemination in 83 regions of Russia (the composition of the Russian Federation at the end of 2013). Arkhangelsk Oblast and Tyumen Oblast are considered separately from the autonomous areas which are their part. EMISS shows zero incidence in the Chechen Republic in 2005 for all diseases. We assume that it is a mistake and that data for this year just are absent. For 2005 there are also not many other data on the Chechen Republic, and data on the monetary income of the population, for example, are available only since 2010 there. Data on drug addiction in every RF subject for 2006 and 2007 in EMISS are the same. Possibly, it is a mistake caused by the fact that in editions of Rosstat data on drug addiction for 2007 are absent. For this reason we took for 2007 average values between 2006 and 2008.

The panel multiple regression equation is the following:

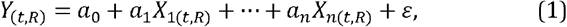

where Y is explained variable, vector with time and space coordinates;

X_i_ – factors affecting the explained variable;

a_i_ – coefficients,

ε – remainder of the equation.

In our case, explained variables – D1-D7 – denote the number of cases or first diagnoses per 100,000 people per year *t* in the region (Russia’s constituent entity) *R, t* takes values from 1 in 2005 to 12 in 2016.

The indicators reflecting the socioeconomic and migration situation, climate and some other characteristics of the region were tested as factors.

Five various indicators of migration were tested in the models.

*The level of legal employment of foreign citizens* (**M1**) is the number of foreign citizens with a valid work permit or a valid employment patent per 1,000 people at the end of a year. This indicator has certain drawbacks. Firstly, data on patents by region are available only since 2013, although patents for foreigners were introduced in the middle of 2010. Secondly, the greatest number of foreign employees in Russia is usually observed in the second and the third quarters, rather than at the end of a year. Thirdly, until 2011, the number of granted permits during a year was published, rather than the number of valid work permits at the end of the year. Thus, the data do not fully meet the comparability criterion.

*The index of international migrant arrival in the region* (**M2**) represents the percentage of arrival in the region coefficient to arrival in Russia coefficient in the given year, i.e. the national level is taken as 1. It is noteworthy that the rules of statistical accounting of international migration for the period 2005–2016 changed many times and even more significantly than statistics on labour migration. As a result, Rosstat data do not reflect the real changes in migration over time, although they help compare regions with each other in each year. So, relative index M2 is more consistent with the criterion of comparability in time and space than arrival coefficient.

Variables M1 and M2 complement each other. Unlike M2, M1 does not include labour migrants from countries of the Eurasian Economic Union (EAEU), but includes migrants who arrive for a period of 3–9 months, while M2 includes migrants arriving for more than 9 months. It can be assumed that illegal migrants are attracted to the same regions as legal ones. Variables M1 and M2 can then be considered to reflect indirectly the impact of illegal labour immigration as well.

*The index of migrant arrival in the region from other RF regions* (**M3**) is calculated by analogy with M2.

*Share of foreign-born population in the region* (**M4**), in percentages.

*Share of population born in other RF regions* (**M5**), in percentages.

Variables M1, M2 and M3 are calculated according to Rosstat data, while M4 and M5 are calculated according to census data of 2002 and 2010.

Four more variables (F_CHN, F_ KAZ, F_GEO and SNR) partly reflect the influence of both migration and geographical factor.

**F_CHN** is a dummy which equals 1 if the region has a common frontier with China, and 0 – if otherwise. Zabaykalsky Krai, Primorsky Krai, Khabarovsk Krai, Altai Republic, Amur Oblast and Jewish Autonomous Oblast share the border with China. We refer Sakhalin Oblast to the group since it is located not far from China oversea.

**F_GEO** is a dummy which equals 1 if the region has a common frontier with Georgia, and 0 – if otherwise. Krasnodar Krai and six republics of North Caucasus Federal District have a common border with Georgia

**F_ KAZ** is dummy which equals 1 if the region has a common frontier with Kazakhstan, and 0 – if otherwise. Astrakhan Oblast, Volgograd Oblast, Kurgan Oblast, Novosibirsk Oblast, Orenburg Oblast, Samara Oblast, Saratov Oblast, Tyumen Oblast, Chelyabinsk Oblast, Altai Republic and Altai Krai have a common frontier with Kazakhstan.

**SNR(Dk)** *is situation with disease ‘k’ in the neighbouring RF regions* (having a common border with the considered region). The average value, weight – migration communications with the neighbouring regions. Migration relations between the region A and region B is the sum of number of natives born in region A living in region B and the number of natives born in region B living in region A. Source of data on migration communications is population censuses of 2002 and 2010. Moscow up to 2012 had the only neighbour Moscow Oblast, since 2012 it also has Kaluga Oblast as a neighbour. Sakhalin Oblast has no land border with other regions; its neighbours oversea are Kamchatka Krai, Primorsky Krai and Khabarovsk Krai. For Kaliningrad Oblast, which is an enclave, value of SNR, equals the average value of incidence in Moscow, St.-Petersburg, Bryansk, Pskov and Smolensk oblasts. Such choice is made on the basis of transport availability and migration relations between the regions. Although when calculating SNR for these five territorial subjects of the Russian Federation, Kaliningrad Oblast was not considered as a neighbour.

The following independent variables were also tested and included in the models, in case of their significance.

*Data on alcohol abuse and alcoholic psychosis diagnosed for the first time, per 10,000 peo*ple (**ALC**). The source is EMISS data.

*Employment rate* (**ER**) is the share of employed among people aged 15–72.

*A number of the Russian tourists sent by tourist firms to foreign trips, per 100,000 people* (**FTT**).

*Hospital Bed-Population Ratio* (**HBPR**), number of available hospital beds for every 10,000 population at the beginning of year.

*Level of education index* in an entity (**ILE**) which is calculated according to census data as the sum of products of share of people (aged 15 years and older) with a certain level of education and the score assigned to this level. Including: no education – 0, primary general education – 1, secondary general and primary vocational education – 3, secondary vocational education –4, incomplete higher education and bachelor’s degree – 5, higher education and postgraduate studies – 6.

*Ratio of monetary income of the population with a living wage*^6^ (**LMI**), i.e. monetary incomes equalling living wage are taken as 1. A set of goods and services in the minimum consumer basket has changed slightly since 2000 therefore it is possible to consider that the LMI variable corresponds to the criterion of comparability.

*Data on living space per person* (**LS**), at the beginning of a year, in sq. meters per person.

*Share of men at the age of 17-19 years, at the beginning of a year* (**M1719**), percentage.

*Dummy* **NatR**, which equals 1 for 26 “national” entities of RF (republics or autonomous entities), and 0 – if otherwise.

*Prevalence HIV infection* (**PHIV**), the share of people living with HIV per 10,000 people. It is calculated according to data from EMISS and the Federal scientific and methodical centre for prevention and fight against AIDS (FCAIDS). When calculating the number of people living with HIV at the beginning of 2006 and the number of the died HIV-positive people in 2005-2014 (according to FCAIDS) as well as number of revealed infected according to EMISS were taken for the basis.

*Population density* (**PD**), people per sq.km.

*Share of pregnant women per 1000 people* (**SPr**), which is calculated using the following formula:

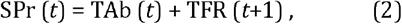

where TAb is number of abortions per 1,000 people ^7^, TFR – total fertility rate per 1,000 people.

*Share of urban population* (**SUP**), at the beginning of a year, percentage.

*Time factor* (***t***) is dummy which equals 1 in 2005, 2 in 2006, …, 12 in 2016.

*The average monthly air temperature in January* (**T1**) and in July (**T7**), according to data of the Federal Service for Hydrometeorology and Environmental Monitoring.

*Share of households without hot water*^8^ (**WHW**), at the beginning of a year, percentage.

*Share of households without sewage*^9^ (**WS**), at the beginning of a year, percentage.

Data source is Rosstat, unless otherwise specified. In case data are calculated on population censuses of 2002 and 2010 (ILE, WHW, WS), it is supposed that values of indicators between censuses changed linearly, and has not changed since 2010.

## 3. Regression analysis and interpretation of results

The following presents the results of econometric modelling by least squares procedure. Equation coefficients are random values, so after each its standard deviation (standard error) is specified in brackets. The significance level of coefficients is indicated by an asterisk: * – 0.1, ** – 0.05, *** – 0.01, **** – 0.001; N – number of observations, R^2^ – determination coefficient. All predictors were included in the models only in case of their significance and in the absence of obvious multicollinearity. It is important as many characteristics of regions are connected with each other. For each period we chose the models with the highest R-squared.

All the models, except for the first year of observation, include incidence in the previous year as a factor. We follow this way since usually the situation with incidence in the region changes slowly. This variable steadily is the most significant for all diseases. It is impossible to include the variable in the model of first-year observation that is why the set of factors in such models differs markedly from the models of following periods. It should be noted that this variable partly reflects the factors operating in the previous year, therefore when this variable is present at the model, other factors, in fact, show characteristics of the regions where the greatest changes happened in comparison with the previous year. Therefore, to complete the picture it is important to consider the dynamics of correlation links of explained variable with various factors while interpreting the results of the regression analysis

### 3.1. HIV

The most significant factors of HIV rate are HIV prevalence rate in the region, intravenous drug use and epidemiological situation in the neighbouring regions. The first predictor is significant in each model presented in table 2, the rest two are significant in all models except for the first one.

**Table 2:** Models to identify new cases of HIV infection

| Models | D1.1 | D1.2 | D1.3 | D1.4 | D1.5 |
| --- | --- | --- | --- | --- | --- |
| Constant | 7.987<br>(8.491) | 41.33****<br>(10.37) | -7.514***<br>(2.427) | 1.552<br>(2.772) | 119.22****<br>(23.60) |
| PHIV | 0.7316****<br>(0.0663) | 0.1859****<br>(0.0320) | 0.5079****<br>(0.0579) | 0.1606**<br>(0.0680) | 0.0758*<br>(0.0417) |
| FTT | 1.375****<br>(0.351) | – | – | – | – |
| Ln(PD) | 3.739****<br>(0.861) | – | – | – | – |
| LMI | 3.547***<br>(1.311) | – | – | – | – |
| T7 | -1.064**<br>(0.403) | – | – | – | – |
| D1(*t*-1) | – | 0.6877****<br>(0.0286) | 0.4087****<br>(0.0558) | 0.7422****<br>(0.0571) | 0.8090****<br>(0.0392) |
| SNR(D1) | – | 0.2003****<br>(0.0189) | 0.1411****<br>(0.0356) | 0.2316****<br>(0.0333) | 0.1192****<br>(0.0310) |
| D4 | – | 0.1626****<br>(0.0470) | 0.2066****<br>(0.0553) | 0.2963***<br>(0.1015) | 0.2004**<br>(0.0845) |
| WS | – | -0.2632****<br>(0.0524) | – | -0.2080***<br>(0.0751) | – |
| ILE | – | -10.55****<br>(2.79) | – | – | -19.86****<br>(5.44) |
| D2 | – | 0.04375***<br>(0.01472) | 0.02956*<br>(0.01686) | – | – |
| NatR | – | -2.010**<br>(0.982) | – | – | – |
| M2 | – | -1.154**<br>(0.554) | – | -2.206**<br>(0.921) | – |
| *t* | – | – | 1.108**<br>(0.538) | – | -3.454****<br>(0.889) |
| WHW | – | – | – | – | -0.2668***<br>(0.0832) |
| N | 82 | 912 | 331 | 332 | 249 |
| R<sup>2</sup> | 0.776 | 0.883 | 0.800 | 0.852 | 0.933 |
| Period | 2005 | 2006-2016 | 2006-2009 | 2010-2013 | 2014-2016 |

**Table 3:** Coverage of HIV-infected with follow-up monitoring and antiretroviral therapy

| Year | Share of HIV-infected under regular medical check-up, from among detected cases | Share of HIV-infected people engaged in antiretroviral therapy, from among HIV-infected under regular medical check-up | Share of HIV-infected people engaged in antiretroviral therapy, from among detected cases |
| --- | --- | --- | --- |
| 2012 | 65,8 | 28,6 | 18,8 |
| 2013 | 64,8 | 32,9 | 21,3 |
| 2014 | 66,0 | 36,4 | 24,0 |
| 2015 | 66,1 | 39,8 | 26,3 |
| 2016 | 72,4 | 42,5 | 30,8 |
| 2017 | 74,2 | 47,8 | 35,5 |

Source: EMISS and FCAIDS

**Table 4:**
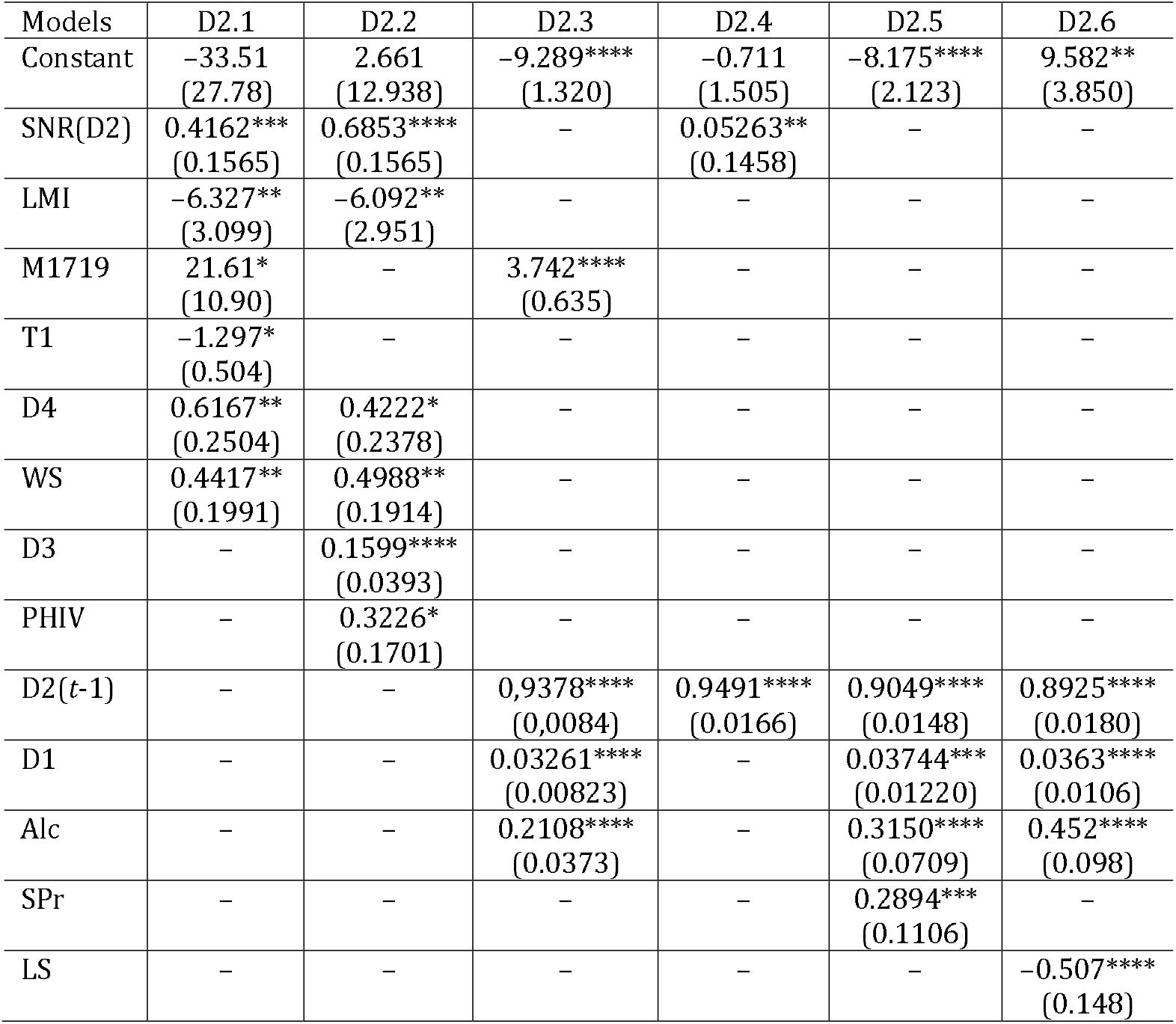
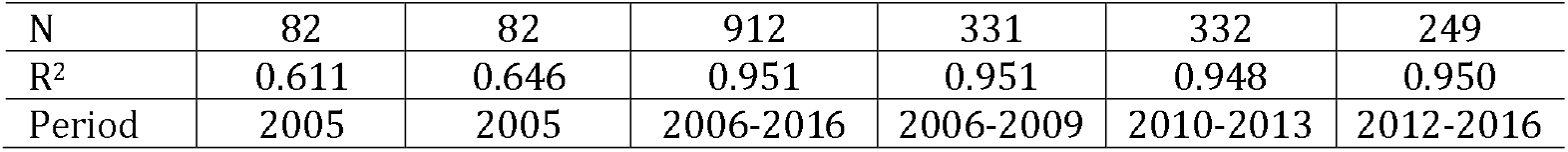
Models to identify new cases of active tuberculosis

| Models | D2.1 | D2.2 | D2.3 | D2.4 | D2.5 | D2.6 |
| --- | --- | --- | --- | --- | --- | --- |
| Constant | -33.51<br>(27.78) | 2.661<br>(12.938) | -9.289****<br>(1.320) | -0.711<br>(1.505) | -8.175****<br>(2.123) | 9.582**<br>(3.850) |
| SNR(D2) | 0.4162***<br>(0.1565) | 0.6853****<br>(0.1565) | - | 0.05263**<br>(0.1458) | - | - |
| LMI | -6.327**<br>(3.099) | -6.092**<br>(2.951) | - | - | - | - |
| M1719 | 21.61*<br>(10.90) | - | 3.742****<br>(0.635) | - | - | - |
| T1 | -1.297*<br>(0.504) | - | - | - | - | - |
| D4 | 0.6167**<br>(0.2504) | 0.4222*<br>(0.2378) | - | - | - | - |
| WS | 0.4417**<br>(0.1991) | 0.4988**<br>(0.1914) | - | - | - | - |
| D3 | - | 0.1599****<br>(0.0393) | - | - | - | - |
| PHIV | - | 0.3226*<br>(0.1701) | - | - | - | - |
| D2(t-1) | - | - | 0.9378****<br>(0.0084) | 0.9491****<br>(0.0166) | 0.9049****<br>(0.0148) | 0.8925****<br>(0.0180) |
| D1 | - | - | 0.03261****<br>(0.00823) | - | 0.03744***<br>(0.01220) | 0.0363****<br>(0.0106) |
| Alc | - | - | 0.2108****<br>(0.0373) | - | 0.3150****<br>(0.0709) | 0.452****<br>(0.098) |
| SPr | - | - | - | - | 0.2894***<br>(0.1106) | - |
| LS | - | - | - | - | - | -0.507****<br>(0.148) |
| N | 82 | 82 | 912 | 331 | 332 | 249 |
| R<sup>2</sup> | 0.611 | 0.646 | 0.951 | 0.951 | 0.948 | 0.950 |
| Period | 2005 | 2005 | 2006-2016 | 2006-2009 | 2010-2013 | 2012-2016 |

**Table 5:** Models to identify new cases of syphilis

| Models | D3.1 | D3.2 | D3.3 | D3.4 | D3.5 |
| --- | --- | --- | --- | --- | --- |
| Constant | -25.02<br>(16.83) | -10.39****<br>(2.38) | -35.42***<br>(12.30) | 7.166<br>(4.664) | -20.07**<br>(7.80) |
| T1 | -2.153***<br>(0.744) | – | – | -0.1829**<br>(0.0823) | – |
| Alc | 2.267****<br>(0.655) | – | – | – | – |
| M1 | 2.223****<br>(0.546) | – | – | – | – |
| D3(t-1) | – | 0.7863****<br>(0.0134) | 0.7929****<br>(0.0234) | 0.7609****<br>(0.0138) | 0.7392****<br>(0.0206) |
| SNR(D3) | – | 0.1385****<br>(0.0270) | 0.1796****<br>(0.0516) | 0.05992**<br>(0.02757) | 0.1050***<br>(0.0381) |
| M1719 | – | 3.946***<br>(1.466) | 11.01**<br>(5.49) | – | – |
| WS | 1.118***<br>(0.362) | 0.0806*<br>(0.0457) | 0.2246*<br>(0.1166) | – | -0.0671*<br>(0.0368) |
| Ln(PD) | – | – | – | 0.6997***<br>(0.2678) | – |
| D5 | – | – | – | 0.04415****<br>(0.01115) | – |
| ER | – | – | – | -0.1714**<br>(0.0752) | – |
| M3 | – | – | – | -1.578***<br>(0.605) | – |
| ILE | – | – | – | – | 5.360***<br>(2.021) |
| F_CHN | – | – | – | – | -2.708**<br>(1.279) |
| N | 82 | 912 | 331 | 332 | 249 |
| R<sup>2</sup> | 0.553 | 0.873 | 0.823 | 0.959 | 0.909 |
| Period | 2005 | 2006-2016 | 2006-2009 | 2010-2013 | 2014-2016 |

**Table 6:** Models to identify new cases of drug addiction

| Models | D4.1 | D4.2 | D4.3 | D3.4 | D3.5 |
| --- | --- | --- | --- | --- | --- |
| Constant | 100.91***<br>(37.89) | 12.52****<br>(2.89) | 18.93****<br>(5.11) | 3.384*<br>(1.714) | 21.24****<br>(3.80) |
| SNR(D4) | 0.4589***<br>(0.1417) | 0.0943****<br>(0.0224) | 0.1152***<br>(0.0366) | – | – |
| ILE | -26.27*<br>(10.46) | -5.315****<br>(0.912) | -8.353****<br>(1.722) | – | – |
| F_GEO | 12.42***<br>(4.57) | – | – | – | – |
| FTT | 1.120***<br>(0.413) | – | – | – | – |
| WHW | -0.2870**<br>(0.1187) | – | – | – | – |
| M5 | 0.3014*<br>(0.1524) |  |  |  |  |
| D4(t-1) | – | 0.7982****<br>(0.0161) | 0.8006****<br>(0.0234) | 0.7762****<br>(0.0253) | 0.9153****<br>(0.0359) |
| SUP | – | 0.0937****<br>(0.0146) | 0.1682****<br>(0.0268) | 0.05582***<br>(0.01655) | – |
| F_CHN | – | 1.773***<br>(0.585) | – | 2.603****<br>(0.635) | 4.271****<br>(1.052) |
| $Ln$(PD) | – | 0.2726***<br>(0.0937) | – | – | – |
| M1 | – | 0.03045**<br>(0.01391) | – | – | – |
| F_Kaz | – | – | – | 1.171**<br>(0.575) | – |
| $t$ | – | – | – | -0.7761****<br>(0.1808) | -1.873****<br>(0.346) |
| N | 82 | 912 | 331 | 332 | 249 |
| R<sup>2</sup> | 0.302 | 0.794 | 0.807 | 0.818 | 0.763 |
| Period | 2005 | 2006-2016 | 2006-2009 | 2010-2013 | 2014-2016 |

**Table 7:** Models to identify new cases of chronic viral hepatitis and acute hepatitis *B* and *C*

| Models | D5.1 | D5.2 | D5.3 | D6 | D7 |
| --- | --- | --- | --- | --- | --- |
| Constant | -132.9***<br>(47.5) | -28.63**<br>(14.07) | -7.063**<br>(3.052) | 0.7725***<br>(0.1949) | -0.6916***<br>(0.2542) |
| SP<sub>r</sub> | 2.971***<br>(0.920) | – | – | – | 0.01402**<br>(0.00697) |
| ER | 2.067***<br>(0.659) | – | – | – | – |
| WS | -1.180***<br>(0.396) | – | – | – | – |
| D3 | 0.2676**<br>(0.1214) | 0.08885****<br>(0.02611) | 0.08873**<br>(0.03633) | – | – |
| M2 | 10.51*<br>(5.38) | – | – | – | 0.06980**<br>(0.03412) |
| D5(*t*-1) | – | 0.8741****<br>(0.0201) | 0.8262****<br>(0.0204) | – | – |
| ILE | – | 8.327**<br>(3.845) | – | – | – |
| LMI | – | – | 2.730***<br>(1.036) | – | 0.1270**<br>(0.0537) |
| ln(PD) | – | – | 0.6313*<br>(0.3289) | 0.03472**<br>(0.01600) | – |
| PHIV | – | – | 0.02823*<br>(0.01711) | – | – |
| D6(*t*-1) | – | – | – | 0.5090****<br>(0.0320) | – |
| NatR | – | – | – | –0.1856***<br>(0.0654) | – |
| D1 | – | – | – | 0.001710**<br>(0.000745) | – |
| *t* | – | – | – | –0.04864***<br>(0.01812) | – |
| D7(*t*-1) | – | – | – | – | 0.5764****<br>(0.0306) |
| D4 | – | – | – | – | 0,008938**<br>(0.003937) |
| F_Kaz | – | – | – | – | 0.2277**<br>(0.0884) |
| SUP | – | – | – | – | 0.004682**<br>(0.002800) |
| N | 83 | 249 | 249 | 498 | 498 |
| R<sup>2</sup> | 0.452 | 0.914 | 0.900 | 0.494 | 0.585 |
| Period | 2010 | 2011-2013 | 2014-2016 | 2011-2016 | 2011-2016 |

Fundamental difference of D.1.1 model from models for the subsequent periods is the importance of FTT predictor. Originally, HIV came to the USSR from abroad, therefore foreign contacts, including trips of Russians, played a role in spread of HIV in Russia. Mainly wealthy residents of large cities had contact with foreigners that is why predictors Ln (PD) and LMI have a plus sign in the model. The correlation of D1 with FTT and Ln (PD) was significant only in the first year of observation (0.364 and 0.351 respectively), and then sharply decreased (in 2014-2016: 0.110 and –0.009)

Further HIV infection spread mainly from the incidence centres created in the territory of Russia therefore SNR (D1) variable is significant in all models the for the subsequent periods.

Though the share of the HIV-positive people who caught through the intravenous use of drugs gradually decreases, the number of incident cases infected through this way keeps growing (fig. 2). In 2014, the number of infected through intravenous drug use (22, 5 thousand people) exceeded the number of incident cases of drug abuse (21 thousand people).

**Fig. 2:**
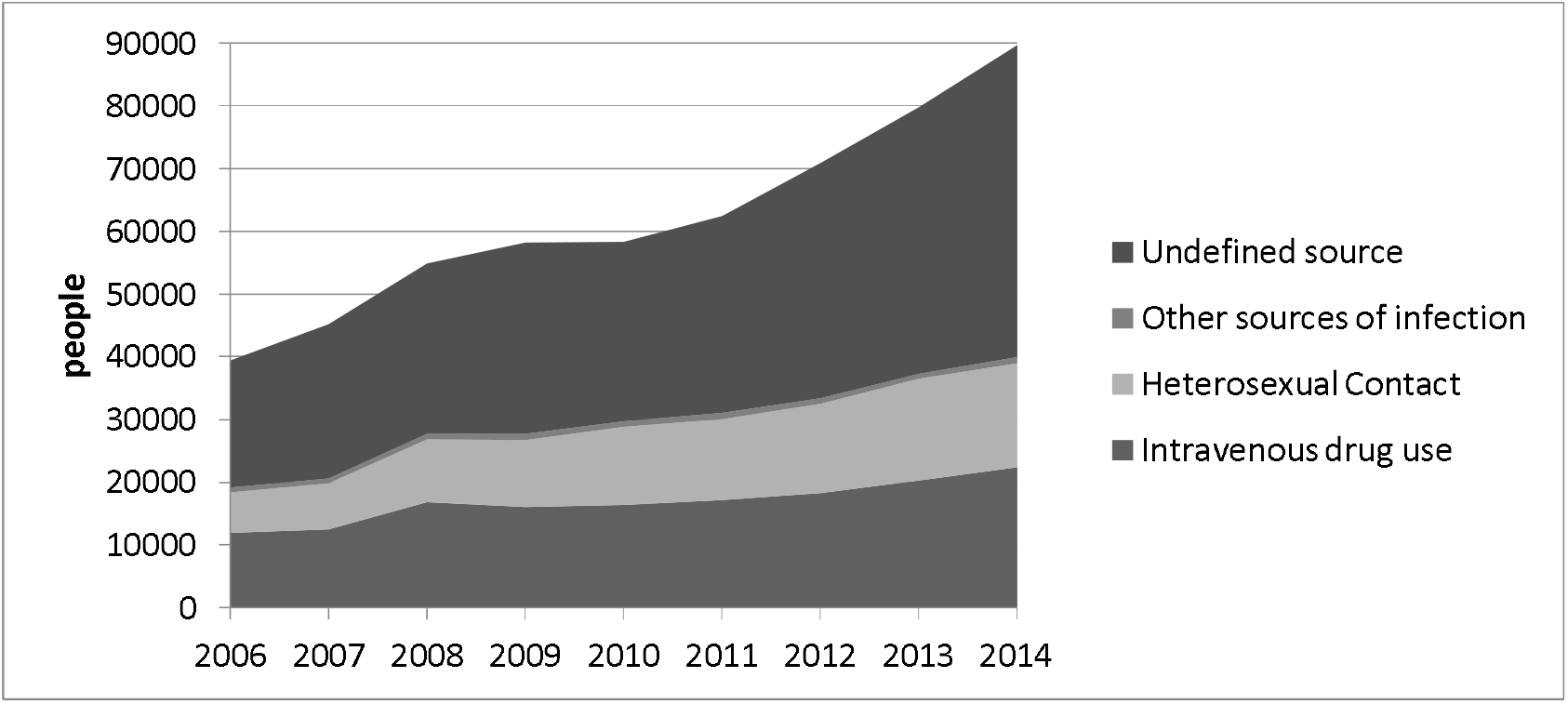
The number of people diagnosed with HIV for the first time among RF residents, by source of infection. Source: HIV infection: information bulletin. No. 40. Central Research Institute of Epidemiology, Federal Research Centre for HIV Prevention and Control. Available at: http://www.hivrussia.ru/files/bul_40.pdf

According to the model for the period of 2006-2016 spread of HIV in “national” regions was slower in coming than in other regions. Traditional features of inhabitants’ behaviour can explain the fact. For example, people quite often refuse to be examined, being afraid to be outcasts.^10^ However, this variable has low priority, and in the models for shorter periods is absent at all.

The main feature of the revealed dynamics of HIV incidence is the increasing distribution of epidemic in less developed regions and, thus, the tendency to align the regions on the incidence of the disease was outlined. The variation coefficient of the indicator of the incidence of HIV decreased from 115.2% in 2005 to 82.1% in 2014-2016. If in the beginning HIV infection extended mainly in economically safe regions, then the situation changed. In models for 2010-2013 and 2014-2016 we can see a combination of “safe” and “unsuccessful” characteristics of the region. Factors WS and WHW with a minus sign in the model are treated as marks of “well-being”. The unattractiveness of the region for migrants (indicator M2 with a minus sign in the model D1.4) as well as an indicator of population education level also entering the model D1.5 with a minus sign are referred to signs of “ill-being”. Thus, eventually indicators of social and economic ill-being amplified among factors of HIV infection spread.

The gradual weakening of correlation link between D1 and some mentioned region characteristics confirms this trend. Thus, the correlation coefficient of D1 with LMI was 0.453 in 2005, 0.267 in 2006-2009 and 0.078 in 2014-2016. The correlation coefficient of D1 with WS was −0.384, −0.234 and −0.146 in the same years; with WHW: −0.415, −0.218 and −0.124; with ILE: 0.487, 0.195 and 0.110 respectively.

The close connection of tuberculosis with HIV infection is well known (Tavares et al. 2017; Farchi et al., 2008). D2 variable is significant in models for the periods 2006-2009 and 2006-2016. Probably, some of TB-infected are also co-infected with HIV. Factor D2 is not significant in D1.4 and D1.5 models, though the correlation coefficient between the level of the revealed HIV incidence, and active tuberculosis grew (from 0.187 in 2006-2009 up to 0.345 in 2014-2016).

According to official data, in 2016 epidemic of HIV is clearly declining, the number of new cases detected significantly decreased, from 68.5 per 100 000 of people in 2015 up to 59.3 in 2016 and 58.4 in 2017.^11^ Therefore, the factor of time is significant with a minus sign in D1.6 model. However, according to experts of the Federal Research Centre for HIV Prevention and Control,^12^ official statistics is incomplete, and incidence actually grew to 69.6 in 2016 and 71.1 per 100 000 of people in 2017.

Providing that official data are correct, then it is possible to assume that the increasing coverage of HIV-infected with antiretroviral therapy pays off and influence of PHIV gradually decreases. However, in actual practice the coverage with antiretroviral therapy remains insufficient (tab. 3) so far, and residents of the regions with the greatest prevalence of HIV, perhaps, prefer to be examined anonymously more often. This can be a reason of difference in data. It is possible also that PHIV role really decreases, but only because the role of other factors increases. Unfortunately, region-wise data of the Federal Research Centre for HIV Prevention and Control are hard to access therefore, it is impossible to construct alternative model for the period 2014-2016 now.

No model of tab. 2 showed the influence of migration on the spread of HIV. Moreover, this is despite the fact that migrants coming to the region have demand for sexual services, while workers of this sphere are risk group.^13^

### 3.2. Tuberculosis

The greatest incidence of tuberculosis is annually observed generally in the same regions, mainly in the Far East and Siberian Federal Districts. Therefore, influence of D2 (t-1) factor in models for incidence of tuberculosis (tab. 4) is higher, than the influence of a similar factor in models for other diseases. The three of the worst in terms of the incidence are Republic of Tyva, Primorsky Krai and Jewish Autonomous Oblast.

According to WHO, the active form of tuberculosis occurs in only a proportion of those infected (from 10% lifetime risk to 10% per year in HIV-positive people) and within a few months or a few years after infection.^14^ HIV infection increases the probability to be infected with tuberculosis and complicates a course of the disease.

Models for tuberculosis show the growing influence of HIV infection. As far as model D2.2 includes PHIV variable, but at the 10% significance level. In models for two last periods when the incidence of HIV significantly grew, the factor D1 is significant at the 1% level. The correlation between D1 and D2 grows eventually. Therefore, the prognosis of tuberculosis depends on how the epidemic of HIV in Russia will progress and how fast the coverage of HIV-positive people with antiretroviral therapy will grow. Such rates as it occurs today (tab. 3), the coverage of 90% of HIV-positive people with antiretroviral therapy will be reached just by 2029.

In addition, other diseases can influence the transition of latent tuberculosis to an active stage.

Links between alcoholism and tuberculosis (TB) incidence is known well (Rudoy et al., 1985; Razvodovsky, 2015). Now it is established that alcohol increases risk of active TB incidence and its development (Molina et al., 2010; Imtiaz et al., 2017). Factor ALC is significant in models D2.5 and D2.6. The correlation between ALC and D2 grew during the considered period, the maximum link was observed in 2014-2016 (0.384).

The reason why drug addiction can influence TB incidence is drug users’ way of life and injury of respiratory tract and decrease in immunity (Silva et al., 2018; Cruz et al., 2013; Kiboi et al., 2016). Factor D4 is significant only in models for 2005, but it is possible to assume, judging by correlation coefficients (from 0.272 in 2006-2009 up to 0.345 in 2014-2016) that further influence of D4 is shown indirectly through D2 (*t*-1).

Question of potential impact of syphilis is disputable (Goldblatt, 1939). That is why, in table 2 we presented two models for 2005, one of them includes D3 as a factor of tuberculosis incidence. This variable is not significant in models for other periods, though the correlation between D2 and D3 is steadily very high (from 0.444 in 2005 up to 0.713 in 2010-2013). It can be explained with the fact that both diseases are most widespread in regions with similar characteristics.

The words of WHO that poverty and poor housing conditions are the main reasons of infectious diseases meet the case of tuberculosis very much. In the Russian realities they can be added by severe climate. The correlation of D2 with T1 (from-0.543 to-0.596), WS (0.337-0.441), LS (from-0.279 to-0.333) and LMI (from-0.195 to-0.308) during the different short periods fluctuates in the small ranges. Factors T1, WS and LMI are significant only in models for 2005, in models for the next years their influence is reflected indirectly, via D2 variable (t-1). LS factor, on the contrary, is significant just in D2.6 model.

There are no significant factors, except for SNR and D2 (t-1) in model for 2006-2009 at all. Probably, factors of tuberculosis incidence during this period were the same as in previous years. The factor SNR emphasizes invariable localization of the centres of the greatest tuberculosis spread, that is in this case it is purely geographical factor, unlike models for HIV where it reflected influence of contacts between people. Correlation of D2 and SNR is steadily very high (0.653-0.696).

During the observation the welfare of the population in general considerably grew. Residential penetration grew for 2005-2015 by 19%, and the rate of increment was quite stable. The indicator of LMI changed less consistently: for 2006-2007 sharp growth by 21.6%, by 2012 a run up to 9.4%, and for 2013-2016 decrease by 12.3%. However, the decrease in tuberculosis incidence in Russia happened only after 2008 and continued until the end of the observation period. Apparently, first of all it concerned children and teenagers therefore the correlation of M1719 with D2 decreases (from 0.499 in 2005 up to 0.189 in 2014-2016). It, obviously, yet did not affect the incidence of the population of other ages, therefore correlation **SPr** and D2 remains steadily high in each span (0.497-0.537).

Pregnant women and young people of military age are categories of the population that are subject to obligatory medical examination. Therefore, the high positive correlation of variables **SPr** and M1719 with incidence can demonstrate both hypo dispersion of the disease on the territory of the country, and problems with detectability of the disease among other categories of the population.

### 3.3. Syphilis

According to models for tuberculosis and HIV, migration had no impact on their spread. Probably the matter is that tuberculosis, HIV and syphilis (unlike viral hepatitis) are included into the list of diseases at which detection the foreign citizen has to leave Russia.^15^

Despite this circumstance, in model for syphilis D3.1 (tab. 5) the inflow of the international labour migrants was a significant factor. It can be explained as follows. Firstly, the incidence of syphilis in the Russian Federation in 2005 was three times higher, than the incidence of HIV. The incidence of tuberculosis was even higher. However, migrants catch sexually transmitted diseases more often after arrival, through the use of sexual services. Secondly, if we assume that illegal labour migrants went to the same regions, as legal ones, the positive importance of M1 factor reflects the circumstance that in 2005 the level of illegal employment of the foreign citizens who did not undergo an obligatory medical check was quite high only in B3.1 model, and then it decreased. The close correlation between D3 and M1 (0.601) was observed only in 2005, in 2006-2009 it fell up to 0.250, and in 2014-2016 up to 0.017.

As well as in a case with tuberculosis, syphilis is more often recorded in cold regions with poor housing conditions. Correlation coefficient of D3 with T1 fluctuated from –0.498 in 2005 up to –0.561 in 2010-2013, and with WS grew: 0.126 and 0.479 in the same years. Unlike tuberculosis, the correlation with alcoholism quickly decreased: from 0.570 in 2005 to –0.027 in 2014-2016.

During 2006-2016 the incidence of syphilis decreased more than by 3 times. However, there is a reason to suppose that such magnificent statistics is a result of underreporting of cases. The fact is pointed by the remaining significant correlation of D3 with M1719 (from 0.348 in 2005 to 0.258 in 2014-2016), high correlation with SPr (from 0.525 in 2005 to 0.575 in 2010-2013) as well as positive significance of Ln (PD) variable in D3.4 model. The latter means that the officially reported incidence rate declined quicker in those places where medical services are less available because of low population density.

Models show that there is an alignment of regions with various welfare on incidence of syphilis over time. During the whole period of observation, the disease was unique to regions that are unsuccessful in social and economic relation. Correlation trend of D3 with WS and LS as well as positive significance of WS in models D3.2 and D3.3 and the negative significance of the LS and M3 in D4 model count in favor of the fact. However, in D3.5 model one can see essentially new phenomenon when ILE enters model signed plus while WS and F_CHN signed minus.

It should be noted that the correlation of D3 with F_CHN during 2006-2016 was significantly positive (0.256-0.411) as all territorial subjects of the Russian Federation bordering China were in top ten regions with the greatest incidence. It is clear, whether it is connected with contacts with China residents. But correlation link of D3 with *Ln* (PD) was, on the contrary, negative though weakened eventually (from –0.494 in 2005 up to –0.195 in 2014-2016). However, there is no multicollinearity in the models as variables *Ln* (PD) in 2010-2013 and F_CHN in 2014-2016 had significant correlation with indicator ΔD3 with those signs with which they entered the corresponding models. Here ΔD3 is a change of the incidence in comparison with previous year:

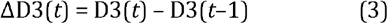

It happened because in 2010-2016 the revealed incidence decreased quicker in those places where its level was the highest while in some safe regions (Moscow, Kaluga Oblast) the incidence even grew.

### 3.4. Drug addiction

Almost all models of new cases of drug addiction identification include fictitious variables reflecting proximity of this or that border (tab. 6). It appears that these variables characterize ways of drugs delivery. As a rule, the Chinese direction dominates, in 2005 it was also Georgia, and in 2010-2013 Kazakhstan. Therefore, there are nidus from where “infection” extends further, therefore in the majority of models the SNR (D4) variable is significant. In this respect drug addiction extension is quite similar to the spread of infections.

Some regions close to China (Primorsky Krai, Sakhalin Oblast, and Jewish Autonomous Oblast) or to Georgia (Krasnodar Krai, the Kabardino-Balkar and Karachay-Cherkess republics) as well as neighbours of these regions (Rep. of Adygea, the Irkutsk and Kemerovo Oblast, Altai and Stavropol Krai) were in 2005 among leaders in the revealed incidence of drug addiction. By the end of the considered period the incidence of drug addiction fell below the average Russian level in the southern regions of the country, in Siberian Federal District it remained above average, and in the Far Eastern Federal District even grew, especially in Primorsky Krai and Jewish Autonomous Oblast.

Primorsky Krai was in the lead in 2005 on the level of foreign trips per 1000 people. Some other regions with high incidence of drug addiction differed also relatively in the large activity in respect of foreign tourism (St. Petersburg, Sverdlovsk Oblast, Jewish Autonomous Oblast, Kabardino-Balkar Republic, Tyumen Oblast without autonomous okrugs, Murmansk and Novosibirsk Oblast, Perm Krai). During the whole considered period the significant positive correlation was between FTT and F_CHN, therefore only FTT in D4.1 model and F_CHN in D4.5 model is significant, according to the fact of what the correlation from these two predictors was higher with explained variable.

Models and correlation links allow to assume that Russian tourists and internal migrants were used as drug traffickers in 2005, later foreign citizens began to prevail among drug couriers. The correlation of D4 with FTT, M5 and M1 was 0.242, 0.232 and –0.026 in 2005, and 0.246, 0.182 and 0.231 in 2014-2016 respectively. However, M1 variable is significant only in model for the long period 2006-2016, but is not significant in short periods.

As a rule, the level of drug addiction is higher in regions with higher population density and in the cities with better housing stock. It, probably, reflects the preferences of drug dealers seeking to get the greatest profit. Quite often families of addicts lost their flats because of his/her debts; especially it was common in the 1990s and at the beginning of the 2000s. In this case, drug mafia was interested in liquid housing equipped with city public services.

Level of education index enters the models D4.1, D4.2 and D4.3 with a minus sign since, as a rule, the higher education level is, the better skills for preventive behavior are developed.

### 3.5. Viral hepatitis

Chronic viral hepatitis and acute hepatitis *C* are mainly revealed in economically safe regions where there is money for examination and treatment of people. High employment rate and sewerage provision of housing act as indicators of well-being in D5.1 model (tab. 7) while education level serve for this purpose in D7.2 model and the monetary income of the population in D5.3 and D7 model. It can be considered the certificate of low detectability of viral hepatitis. The version is also proved by the importance of variable SPr in D5.1 and D7 models and growth of correlation between D5 and SPr (from 0.173 in 2005 to 0.274 in 2011-2016). The significance of population density and share of urban population with a plus sign in D6 and D7 models tells about the same: detection of the disease is lower in the places where medical care is less available.

Factor M2, the arrival of the international migrants in the region, is significant in D5.1 and D7 models. It is not surprising since the prevalence of viral hepatitis in the majority of the countries of origin is higher, than in the Russian Federation.

At the same time the correlation of M2 with D5 and D7 considerably decreases (for D5: from 0.374 in 2010 to 0.179 in 2014-2016, for D7: from 0.305 in 2011-2013 up to 0.225 in 2014-2016).

D6 model includes NatR variable with a minus sign. It is not clear, whether it follows from the fact that hepatitis *B* virus is spread less in the majority of “national” regions, or people are less examined, as a rule, for this disease there.

The reason of variable F_ Kaz significance in D7 model is also not quite clear. It can be connected either with migration from Kazakhstan or with better-organized screening for hepatitis *C* in the Ural Federal District. Some regions which share border with Kazakhstan (Kurgan, Tyumen and Chelyabinsk oblasts) or have high share of natives of Kazakhstan (Sverdlovsk oblast, Yamalo-Nenets and Autonomous Okrugs and Khanty-Mansiisk autonomous okrug) as a part of resident population are among regions with high incidence of acute hepatitis *C*, but others are not. The variable characterizing a share of natives of Kazakhstan in the population (according to thr census of 2010), was tested and was not significant.

The models showed high correlation between viral hepatitis and other diseases considered above. D7 model tells about drug addiction as hepatitis *C* spreading factor. There is linkage between chronic viral hepatitis and syphilis: D5 variable is significant in D3.4 model, D3 variable is significant in D5 model. Probably, people infected with one sexually transmitted disease are also checked for other diseases. However, the correlation between syphilis and acute hepatitis *B* and *C* is absent. The links between viral hepatitis and HIV is also observed: D1 factor is significant in D6 model, PHIV factor – in D5.3 model. Correlation linkage PHIV and D5 amplifies: from 0.137 in 2010 to 0.258 in 2014-2016. It is possible, for the reason, that HIV increases risk of transition of acute hepatitis *B* and *C* to chronic form.

For each acute viral hepatitis only one model is done since there are not enough registered cases of diseases. It can be connected with very low detectability of these diseases.

The fact that new cases of chronic viral hepatitis are registered several times more, than acute hepatitis *B* and *C* indirectly testifies it.

## 4. Conclusion

The conducted research allows us to conclude that the incidence of a number of socially dangerous diseases (tuberculosis and syphilis) is closely related to the general unsatisfactory economic and social situation in the region (poverty, poor housing conditions, alcoholism, and low employment rate). Some others (HIV infection and viral hepatitis), on the contrary, are more widespread in economically successful regions. Perhaps, there are more problems with detection of these diseases in poor regions. Drug addiction holds special position: links to socioeconomic indexes are minimized, while the models include variables reflecting drug trafficking and convenience of drug distribution for dealers. In this case proximity to borders (firstly to Georgia, then to China and Kazakhstan) as well as migration, both internal and external, play an important role.

The research allowed us to establish positive significant statistical correlation of the following indicators of migration and incidence: employment of foreign citizens and incidence of syphilis in 2005; share of internal migrants in the population and incidence of drug addiction in 2005; employment of foreign citizens and incidence of drug addiction in 2006-2016; inflow of foreign citizens and detection of chronic viral hepatitis in 2010; inflow of foreigners and detection of acute hepatitis *C* in 20011-2016.

Reduction of illegal employment of foreign citizens from the visa-free countries through easing administrative barriers to legal employment allowed to reduce the probability of international migration influence on the spread of infectious diseases. However, there is a problem with receiving by migrants the money that they earned as the state does not protect the right of foreign workers if the employer involves them without the conclusion of the official employment contract. As a result the state pushes migrants to illegal work, including the prohibited fields of activity, for example, as drug traffickers.

The research proved that drug addiction has impacted on the spread of HIV and virus of hepatitis *C*, as well as increases the probability to be infected with tuberculosis. Thus, we can say, that indirectly illegal migration promotes spread of these diseases. Therefore, protection of labour rights of the international migrants by the state could promote decreases in incidence of Russian citizens.

## Data Availability

Data openly available within the article and in public repository that does not issue DOIs

http://www.euro.who.int/en/health-topics/health-determinants/migration-and-health/migrant-health-in-the-european-region/migration-and-health-key-issues

http://www.gks.ru/free_doc/new_site/population/urov/met_2.htm

https://www.svoboda.org/a/29281743.html

http://aids-centr.perm.ru/images/4/hiv_in_russia/hiv_in_rf_31.12.2017.pdf

http://www.hivrussia.ru/files/bul_40.pdf

## Footnotes

1 WHO regional office for Europe. Migration and Health: key issues. Available at: http://www.euro.who.int/en/health-topics/health-determinants/migration-and-health/migrant-health-in-the-european-region/migration-and-health-key-issues (accessed: 04.08.2019).

2 Approved by the Resolution of the Government of the Russian Federation no. 715, dated 01.12.2004. Available at: http://base.garant.ru/12137881/ (accessed: 19.03.2018).

3 Unified Interdepartmental Statistical Information System. Available at: https://fedstat.ru/organizations/ (accessed: 01.07.2019).

4 The Ministry of Health provides statistics separately on “active tuberculosis” and “tuberculosis of respiratory tract”. In the List of socially significant diseases at Rosstat website only “active tuberculosis” is listed, although the List approved by the Government of the Russian Federation contains “tuberculosis” without specification.

5 diagnosed for the first time; the virus is not specified (hepatitis *B* and *C* can become chronic)

6 The living wage is the conditional minimum income necessary for ensuring the minimum basic needs of the person. It includes a conditional set of food products (50% of the minimum consumer basket), nonfoods and services (50% of the minimum consumer basket) and obligatory tax payments (13% of income, only for able-bodied population, taking into account tax deduction on children). The size of a living wage in regions is established by authorities of territorial subjects of the Russian Federation on the basis of goods prices and service in the previous quarter (separately for children, able-bodied population and pensioners and also average for the population of the region). The size of a living wage is the cornerstone of calculations for social payments. More information is available at: http://www.gks.ru/free_doc/new_site/population/urov/met_2.htm.

7 In 2006-2016 TAb values decreased quickly enough. In 2006 there were 96 abortions for 100 born children, in 2016 the number of abortions accounted for 39 abortions of 100 born children.

8 There is neither centralized hot water service, no individual water heaters. According to census-2002, the share of such households in the Russian Federation was 39.6%. In 29 territorial subjects of the Russian Federation from 83 considered a share of households without hot water was more than 50%. According to census of 2010 the share of such households in the Russian Federation was 25.4%, from 0.56% in Moscow up to 77.2% in Altai Republic.

9 The toilet is outhouse or absent. According to census-2002, the share of the households without sanitary sewer was 33.9%, according to census −2010 – 17.0% (from 0.06% in Moscow up to 65.3% in the Republic of Tyva). One of the reasons for gradual decrease in WHW and WS is departure or death of inhabitants. Between censuses of 2002 and 2010 the number of the RF settlements having at least one permanent resident decreased from 145143 to 136072.

10 Sourse: https://www.svoboda.org/a/29281743.html

11 https://www.rosminzdrav.ru/news/2018/11/29/9929-sovmestnyy-kommentariy-minzdrava-rossii-i-rospotrebnadzora-o-zabolevaemosti-vich-spidom-v-rossii

12 http://aids-centr.perm.ru/images/4/hiv_in_russia/hiv_in_rf_31.12.2017.pdf

13 In 2017 selective examination of commercial sex workers in four large cities of the Russian Federation was conducted. 2.3% of sex workers were HIV infected in in St. Petersburg, 5.4% in Krasnoyarsk, 14.2% in Yekaterinburg and 15.0% in Perm. It is many times less, than prevalence of HIV infection among injecting drug users: 48.3%, 48.1%, 57.2% and 64.6% in the same cities respectively. Source: http://aids-centr.perm.ru/images/4/hiv_in_russia/hiv_in_rf_31.12.2017.pdf For comparison: at the date of 31.12.2014 the greatest established prevalence of HIV was observed in men 30-34 years (2.37%), men 35-39 years (2.28%) and women 30-34 years (1.79%), other gender and age groups have less prevalence. Source: http://www.hivrussia.ru/files/bul_40.pdf

14 WHO regional office for Europe. Migration and Health: key issues. Available at: http://www.euro.who.int/en/health-topics/health-determinants/migration-and-health/migrant-health-in-the-european-region/migration-and-health-key-issues

15 The list of Infectious socially dangerous diseases justifying the refusal to issue or cancellation of the temporary residence permit of foreign citizens and stateless persons, or residence permit, or patent, or work permit in Russia: Order of the Ministry of Health of the Russian Federation No. 384n, dated 29.06.2015.

